# A systematic review and meta-analysis of published research data on COVID-19 infection-fatality rates

**DOI:** 10.1101/2020.05.03.20089854

**Authors:** Gideon Meyerowitz-Katz, Lea Merone

**Affiliations:** Western Sydney Local Health District; University of Wollongong; James Cook University; Apunipima Cape York Health Council

## Abstract

An important unknown during the COVID-19 pandemic has been the infection-fatality rate (IFR). This differs from the case-fatality rate (CFR) as an estimate of the number of deaths as a proportion of the total number of cases, including those who are mild and asymptomatic. While the CFR is extremely valuable for experts, IFR is increasingly being called for by policy-makers and the lay public as an estimate of the overall mortality from COVID-19.

**Methods:** Pubmed, Medline, SSRN, and Medrxiv were searched using a set of terms and Boolean operators on 25/04/2020 and re-searched 14/05/2020, 21/05/2020, and 16/06/2020. Articles were screened for inclusion by both authors. Meta-analysis was performed in Stata 15.1 using the metan command, based on IFR and confidence intervals extracted from each study. Google/Google Scholar was used to assess the grey literature relating to government reports.

**Results:** After exclusions, there were 24 estimates of IFR included in the final meta-analysis, from a wide range of countries, published between February and June 2020.

The meta-analysis demonstrated a point-estimate of IFR of 0.68% (0.53-0.82%) with high heterogeneity (p<0.001).

**Conclusion:** Based on a systematic review and meta-analysis of published evidence on COVID-19 until May, 2020, the IFR of the disease across populations is 0.68% (0.53-0.82%). However, due to very high heterogeneity in the meta-analysis, it is difficult to know if this represents the ‘true’ point estimate. It is likely that, due to age and perhaps underlying comorbidities in the population, different places will experience different IFRs due to the disease. Given issues with mortality recording, it is also likely that this represents an underestimate of the true IFR figure. More research looking at age-stratified IFR is urgently needed to inform policy-making on this front.

**Key messages:**

– COVID-19 infection-fatality rate (IFR) is an important statistic for policy about the disease
– Published estimates vary, with a ‘true’ fatality rate hard to calculate
– Systematically reviewing the literature and meta-analyzing the results shows an IFR of 0.68% (0.53-0.82%)

## Introduction

2020 saw the emergence of a global pandemic, COVID-19, caused by the SARS-CoV-2 virus, which began in China and has since spread across the world. One of the most challenging questions to answer during the COVID-19 pandemic has been regarding the true infection-fatality rate (IFR) of the disease. While case-fatality rates (CFR) are eminently calculable from various published data sources (1) – CFR being the number of deaths divided by the number of confirmed cases - it is far more difficult to extrapolate to the proportion of all infected individuals who have died due to the infection because those who have very mild, atypical or asymptomatic disease are frequently left undetected and therefore omitted from fatality-rate calculations (2). Given the issues with obtaining accurate estimates, it is not unexpected that there are wide disparities in the published estimates of infection. This is an issue for several reasons, most importantly in that policy is dependent on modelling, and modelling is dependent on assumptions. If we do not have a robust estimate of IFR, it is challenging to make predictions about the true impact of COVID-19 in any given susceptible population, which may stymie policy development and may have serious consequences for decision-making into the future. While CFR is a more commonly-used statistic, and is very widely understood among experts, IFR provides important context for policy makers that is hard to convey, particularly given the wide variation in CFR estimates. While CFR is naturally a function of the denominator – i.e. how many people have been tested for the disease – policy-makers are often most interested in the total burden in the population rather than the biased estimates given from testing only the acutely unwell patients.

This is particularly important when considering the reopening of countries post ‘lockdown’. Depending on the severity of the disease, it may be reasonable to reopen services such as schools, bars, and clubs, at different timings. Another salient point is the expected burden of disease in younger age groups – while there are likely long-term impacts other than death, it will be important for future planning to know how many people in various age groups are likely to die if the infection becomes widespread across societies. Age-stratified estimates are also important as it may give countries some way to predict the number of deaths expected given their demographic breakdown.

There are a number of methods for investigating the IFR in a population. Retrospective modelling studies of influenza have successfully predicted the true number of cases and deaths from influenza-like-illness records and excess mortality estimates (3, 4). However, these may not be accurate, in part due to the general difficulty in attributing influenza cases to subsequent mortality, meaning that CFRs may both overestimate and equally underestimate the true number of deaths due to the disease in a population (5). The standard test for COVID-19 involves polymerase chain reaction testing (PCR) of nasopharyngeal swabs from patients suspected of having contracted the virus. This can produce some false negatives (6), with one study demonstrating almost a quarter of patients experiencing a positive result following up to two previous false negatives (7). PCR is also limited in that it cannot test for previous infection. Serology testing is more invasive, requiring a blood sample, however it can determine if there has been previous infection and can be performed rapidly at the point of care (PoC). Serology PoC testing cannot determine if a person is infectious, or if infection is recent and there is risk of misinterpretation of results (8). Serology testing is more sensitive and specific than PCR, but will still likely overestimate prevalence when few people have been infected with COVID-19 and underestimate in populations with more infections (9).

Given the emergence of COVID-19 as a global pandemic, it is somewhat unlikely that these issues are directly mirrored for the newer disease, but there are likely similarities between the two. Some analysis in mainstream media publications and pre-prints has implied that there is a large burden of deaths that remains unattributed to COVID-19. Similarly, serological surveys have demonstrated that there is a large proportion of cases that have not been captured in the case numbers reported in the U.S., Europe, and potentially worldwide (10-12).

This paper presents a systematic effort to collate and aggregate these disparate estimates of IFR using an easily replicable method. While any meta-analysis is only as reliable as the quality of included studies, this will at least put a realistic estimate to the IFR given current published evidence.

## Methods

This study used a simple systematic review protocol. PubMed, MedLine, and Medrxiv were searched on the 25/04/2020 using the terms and Boolean operators: (infection fatality rate OR ifr OR seroprevalence) AND (COVID-19 OR SARS-CoV-2). This search was repeated on 14/05/2020, 25/05/2020, and 16/06/2020. The preprint server SSRN was also searched on 25/05/2020, however as it does not allow this format the Boolean operators and brackets were removed. While Medrxiv and SSRN would usually be excluded from systematic review, given that the papers included are not peer-reviewed, during the pandemic it has been an important source of information and contains many of the most recent estimates for epidemiological information about COVID-19. Inclusion criteria for the studies were:

– Regarding COVID-19/SARS-CoV-2 (i.e. not SARS-CoV-1 extrapolations)
– Presented an estimated population infection-fatality rate (or allowed calculation of such from publicly-available data)

Titles and abstracts were screened for eligibility and discarded if they did not meet the inclusion criteria. GMK then conducted a simple Google and Google scholar search using the same terms to assess the grey literature, in particular published estimates from government agencies that may not appear on formal academic databases. LM assessed the articles to ensure congruence. If these met the inclusion criteria, they were included in the systematic review and meta-analysis. Similarly, Twitter searches were performed using similar search terms to assess the evidence available on social media. Estimates for IFR and the confidence interval were extracted for each study.

All analysis and data transformation was performed in Stata 15.1. The meta-analysis was performed using the metan command for continuous estimates, with IFR and the lower/upper bounds of the confidence interval as the variables entered. This model used the DerSimonian and Laird random-effects method. The metan command in Stata automatically generates an I2 statistic that was used to investigate heterogeneity. Histograms were visually inspected to ensure that there was no significant positive or negative skew to the results that would invalidate this methodology. For the studies where no confidence interval was provided, one was calculated.

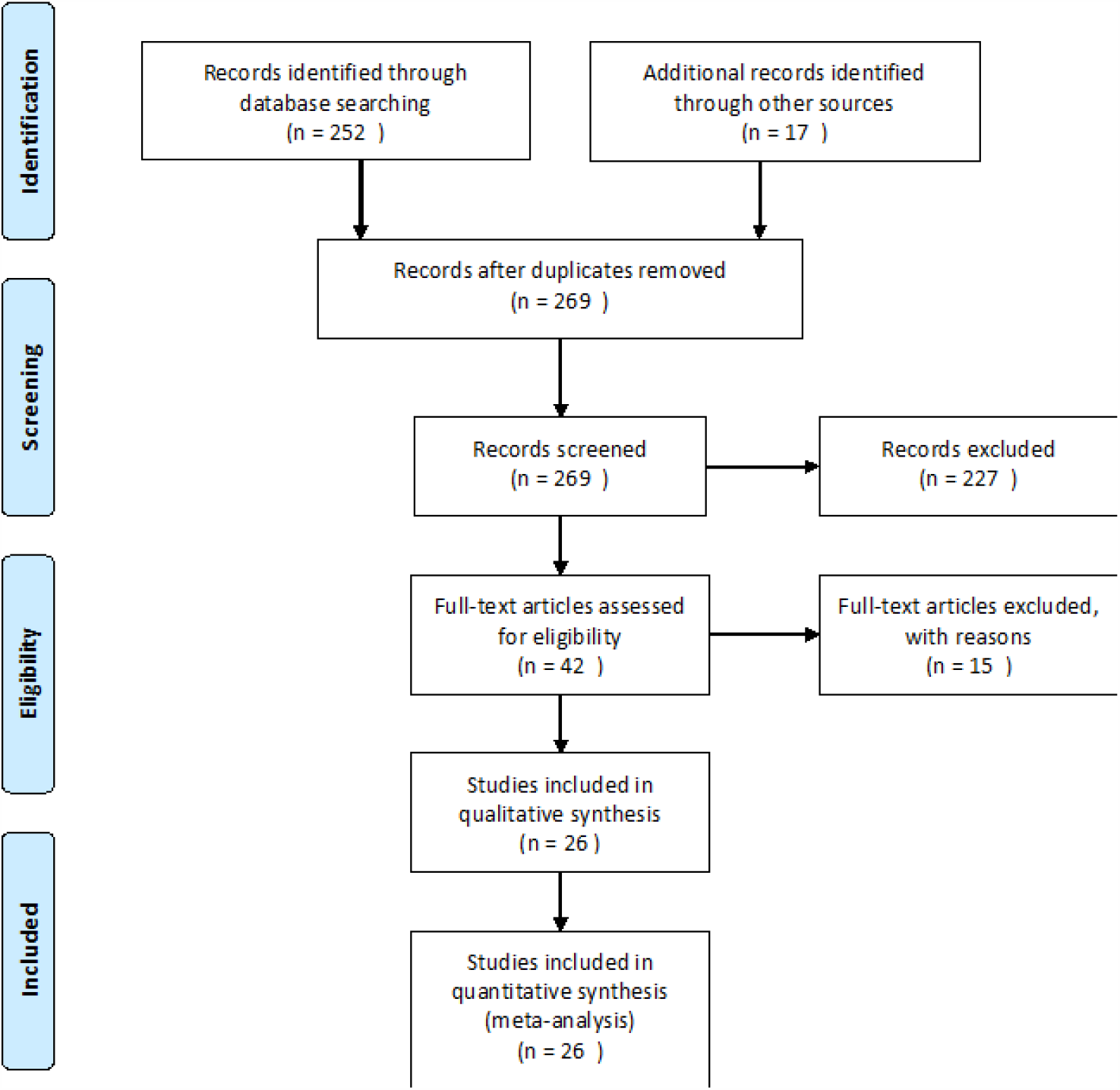

A PRISMA flow diagram of the search methods

Sensitivity analyses were performed stratifying the results into the type of study – serological vs non – by country, and by month of calculation.

The metabias and metafunnel commands were used to examine publication bias in the included research, with Egger’s test used for the metabias estimation. It was challenging to formally rate the risk of bias of the included modelling studies, as there was very significant heterogeneity in methodology and implementation, with the result that the risk of bias in these studies was considered to be high across all included research. Serological surveys were rated using a the risk of bias in prevalence tool with a resulting estimate in line with Cochrane GRADE criteria of low, moderate, or high (13). This tool asks a series of 10 questions about the sampling and data collection of prevalence studies, with a final rating based on the previous questions. Each question is answered yes/no, with a lack of information presumed to be no/unclear. A separate sensitivity analysis was conducted using only serological survey results stratified by risk of bias.

Due to a recent surge in the number of serological surveys being published, these were included in the infection-fatality estimate despite not formally calculating an IFR in the study text itself. Regional death rates were taken from the John Hopkins University CSSE dashboard (14) 10 days after the serosurvey completion where no IFR was calculated to account for right-censoring of these estimates (15), and used to estimate the IFR given the population.

All code and data files are available (in .do and .csv format) upon request.

## Results

Initial searches identified 252 studies across all databases. Later searches on Google and social media, as well as resampling the included databases revealed a further seventeen estimates to include in the study. These came from a variety of sources, with some appearing from blog posts, others posted on Twitter, and some government documents being found through Google. There were no duplicates specifically, however two pre-prints had been published and so appeared in slightly different forms in both databases. In this case, the published study was used rather than the pre-print. Results are collated in table 1.

**Table 1:**
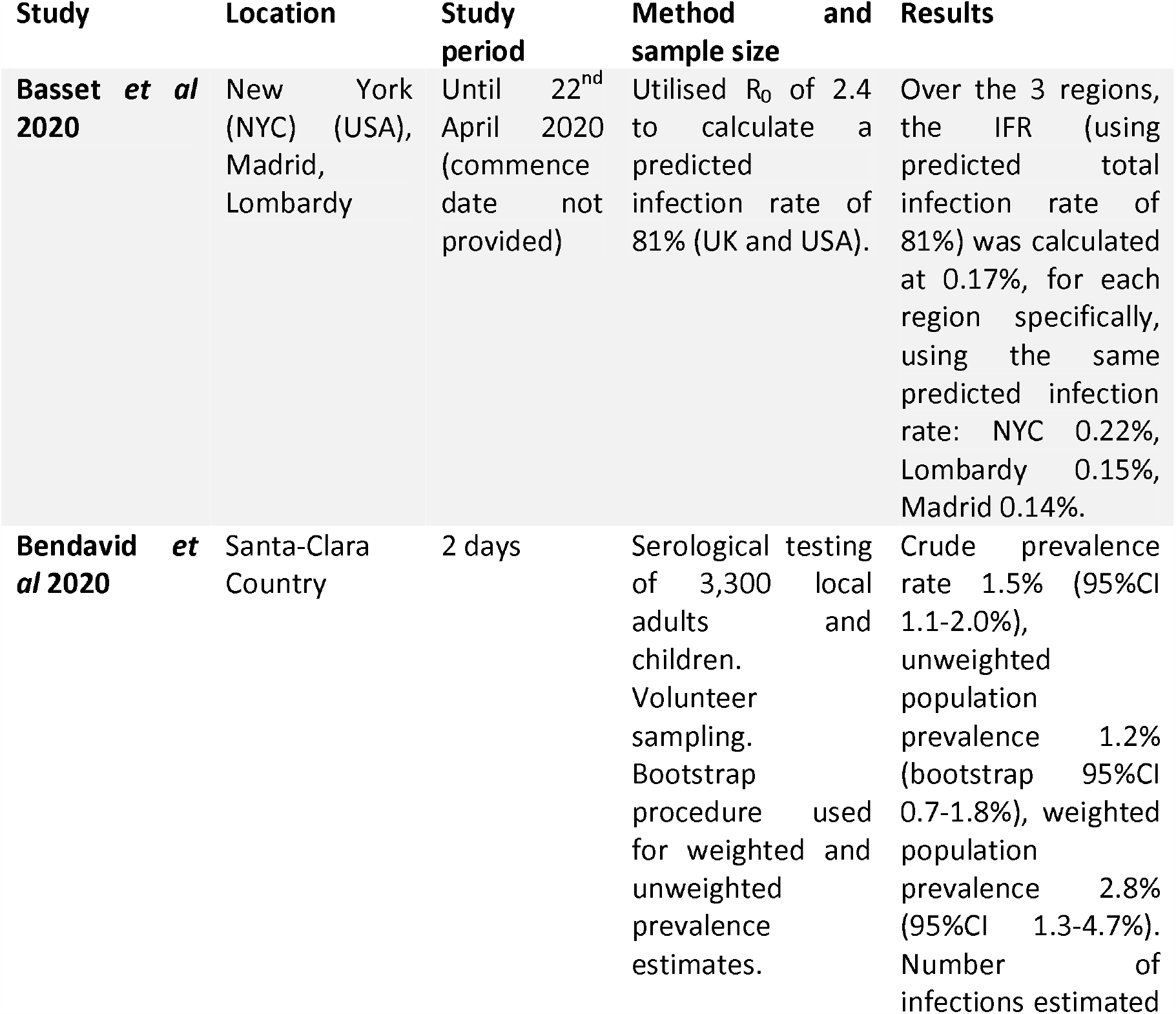
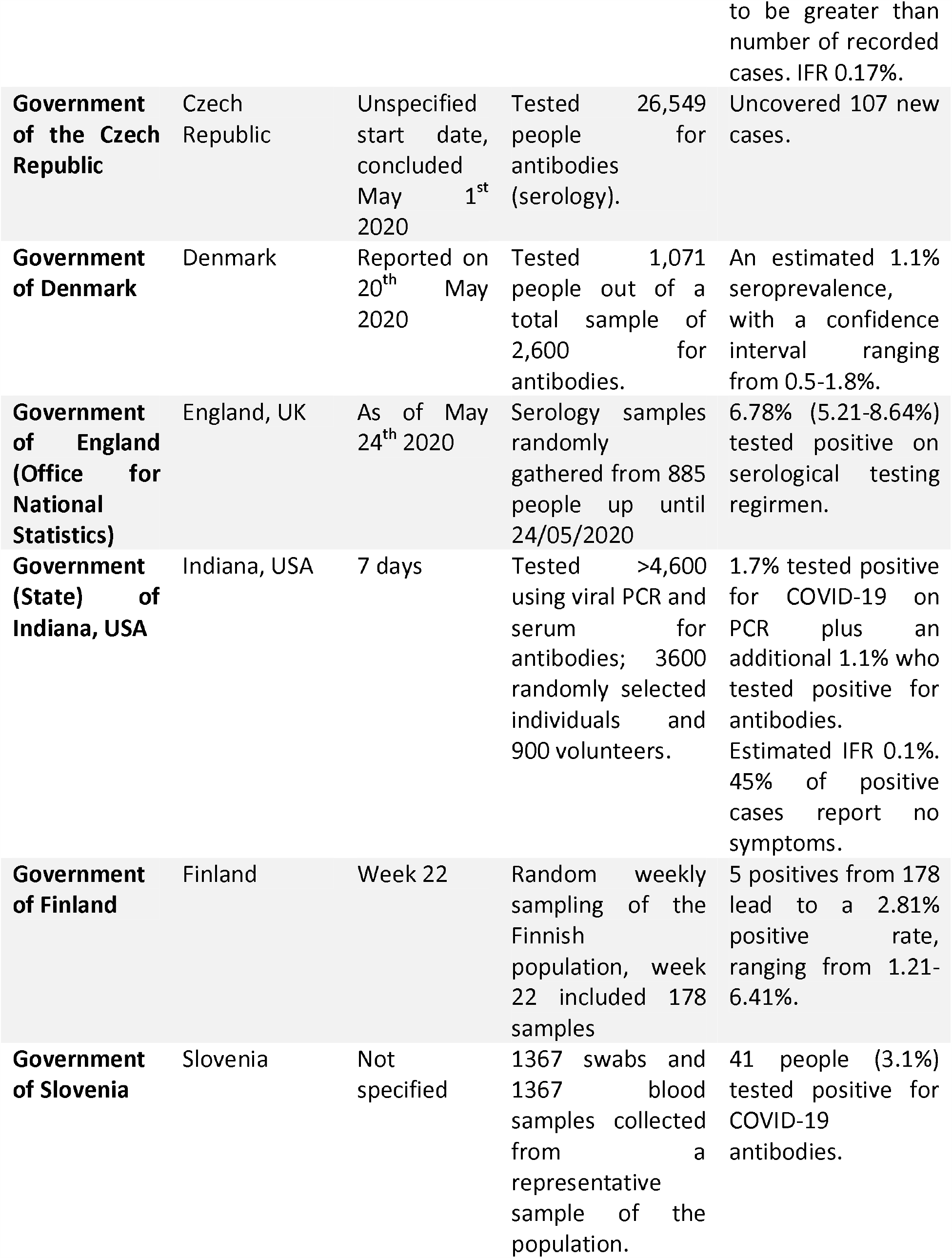
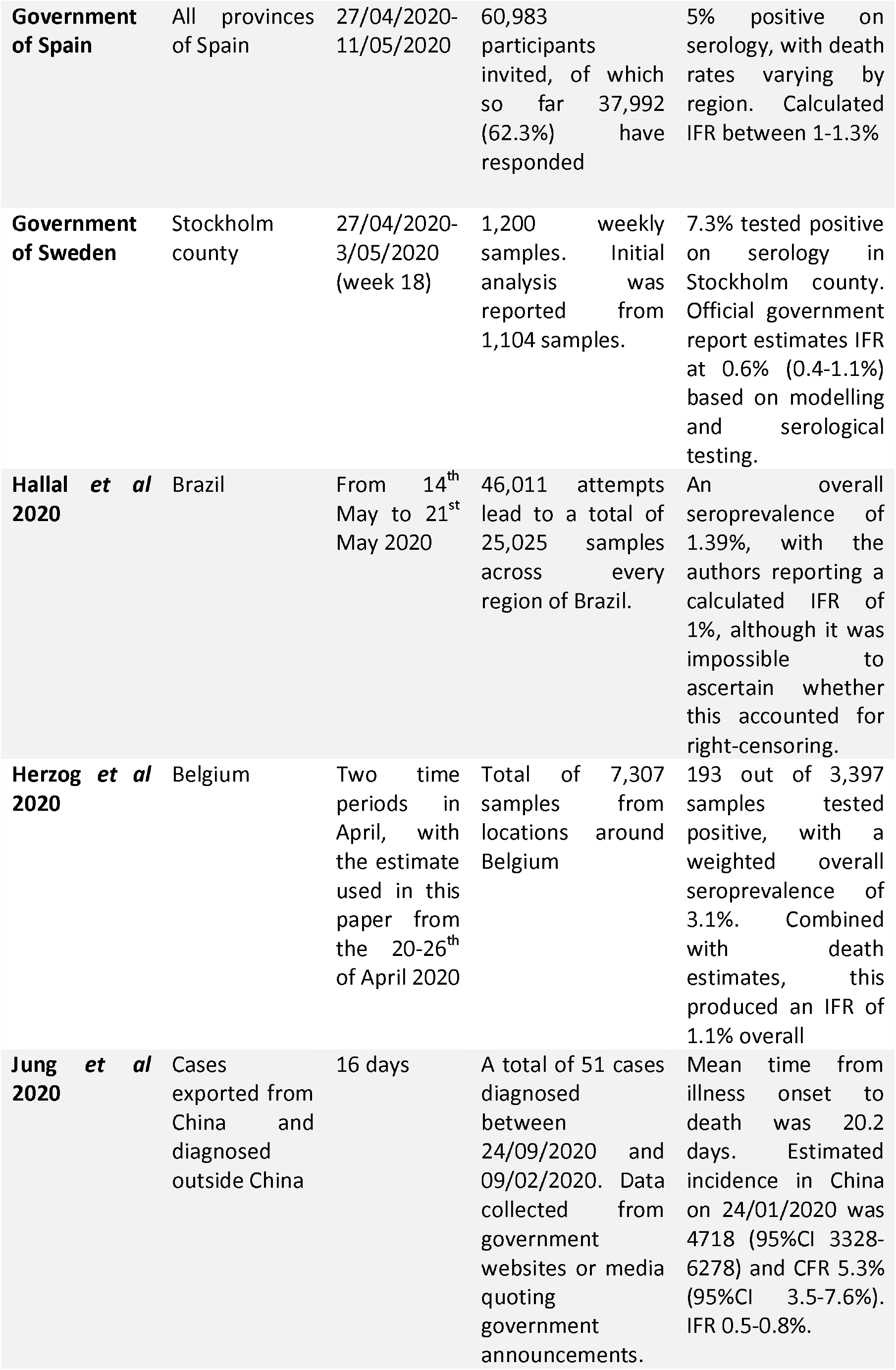
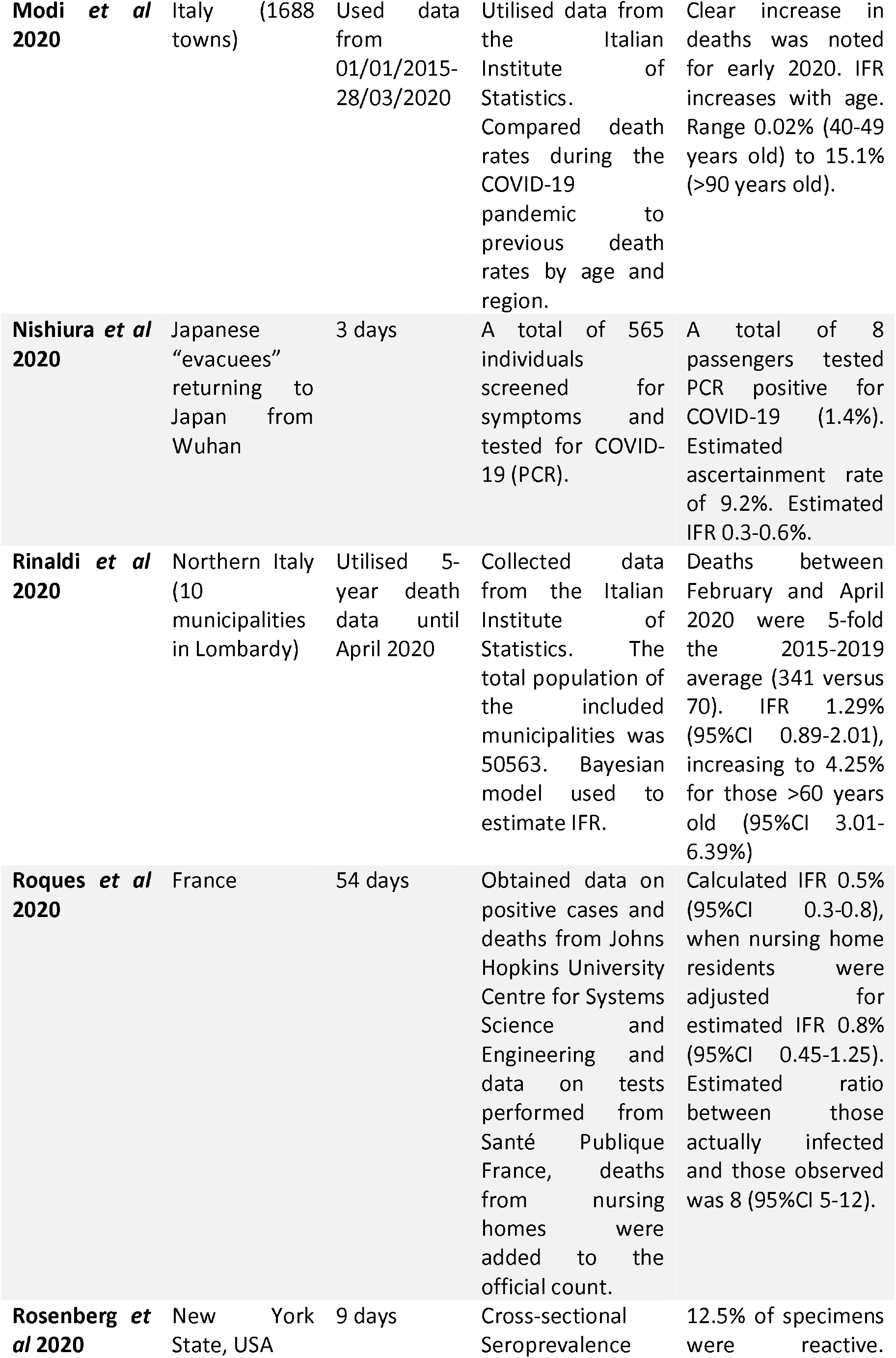
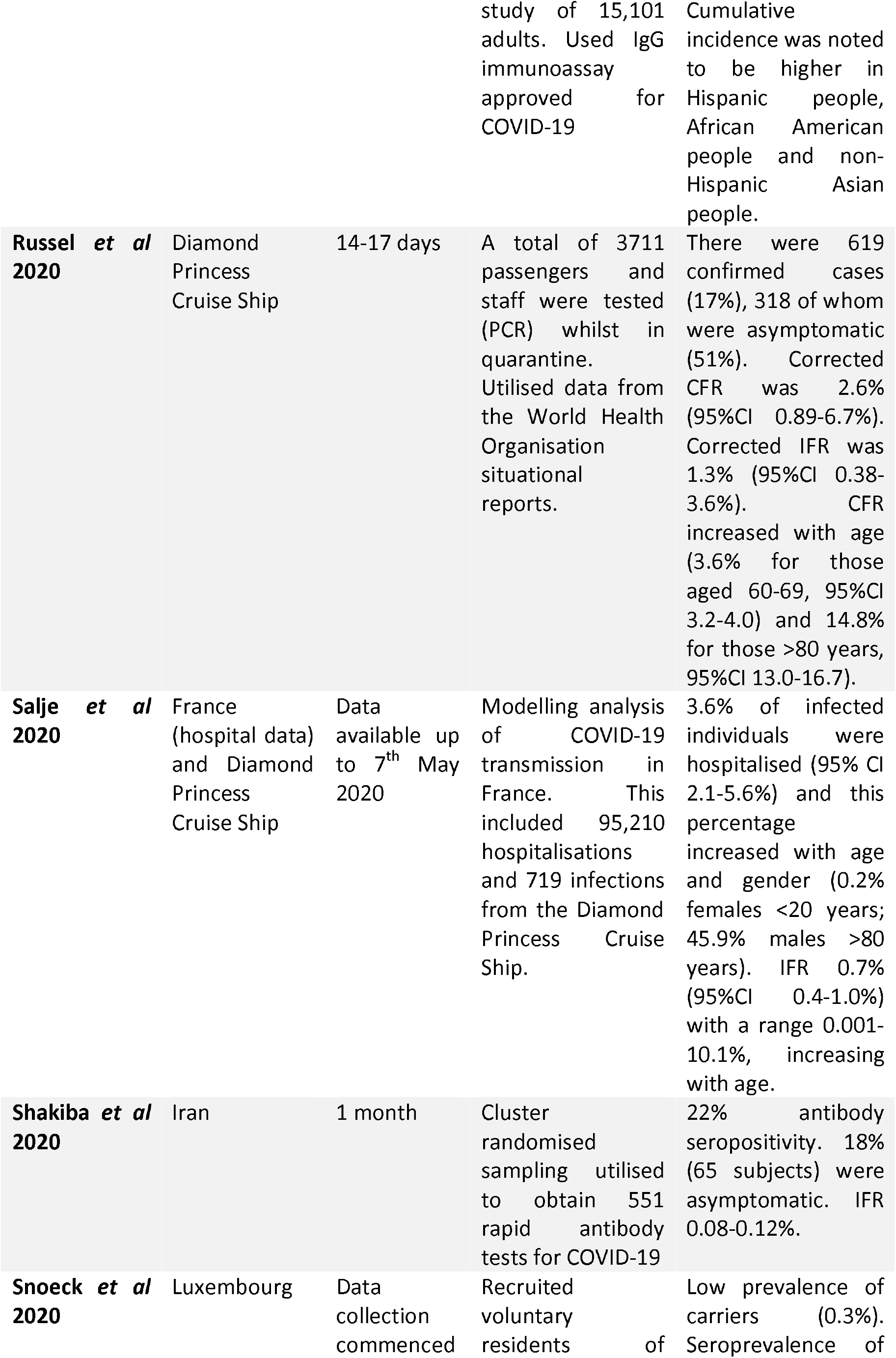
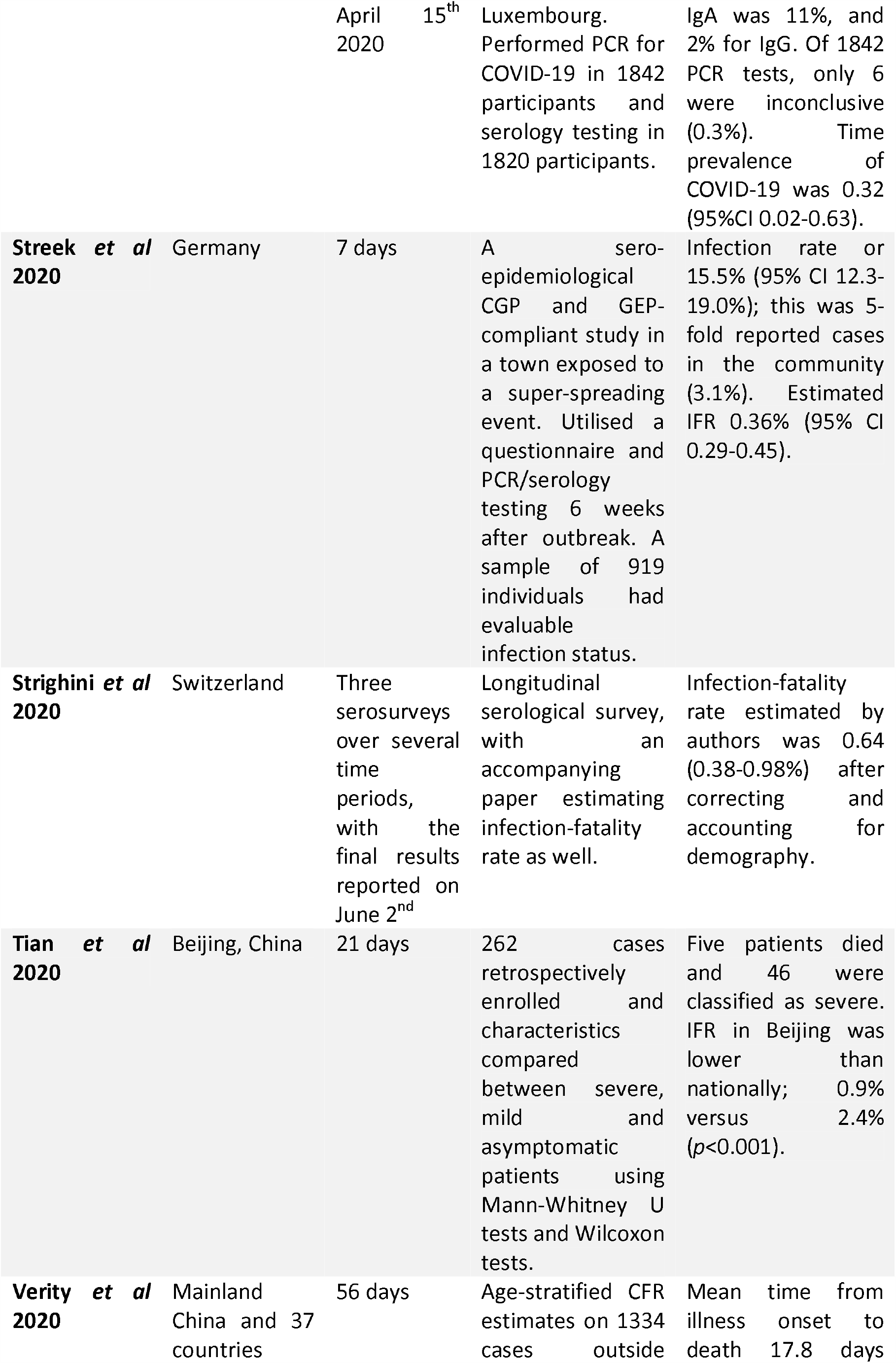
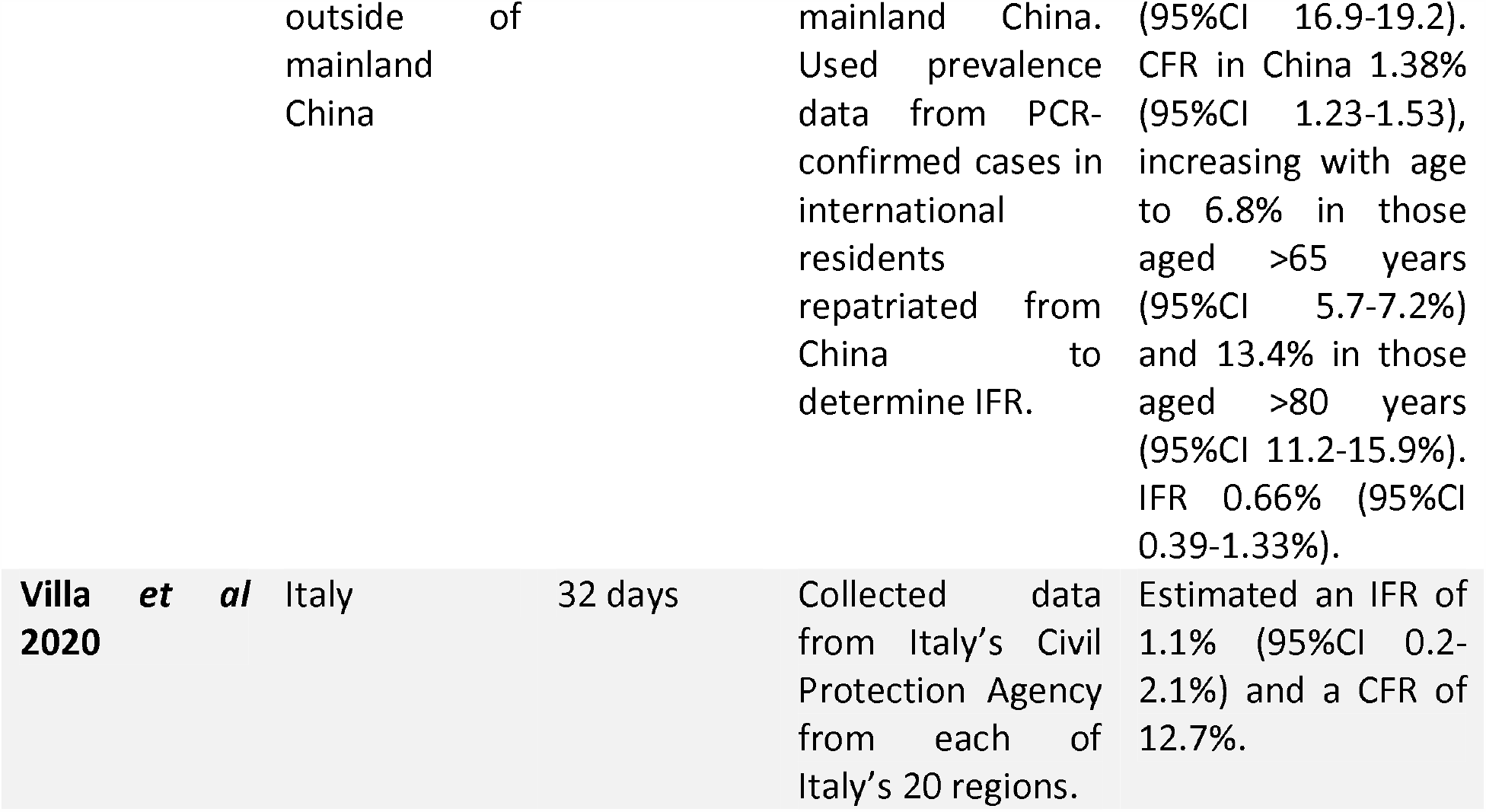
**number: Results of systematic review of published research data on COVID-19 infection-fatality rates**

**Table 2 –.** risk of bias in included serosurveys

| **Study** | **Overall Risk of Bias** |
| --- | --- |
| New York City | Moderate |
| Bendavid et al | High |
| Streeck et al | Low |
| Spain | Low |
| Indiana | High |
| Shakiba et al | Moderate |
| Sweden (Stockholm) | Moderate |
| Stringhini et al | Low |
| Wu et al | High |
| Snoeck et al | Low |
| Slovenia | High |
| Czech Republic | Low |
| Denmark | Moderate |
| Hallal Et al | Low |
| Herzog et al | Moderate |
| Finland | Moderate |
| ONS England | Moderate |

Studies were excluded for a variety of reasons. Some studies only looked at COVID-19 incidence, rather than prevalence of antibodies, and were thus considered potentially unreliable as population estimates (16). The most common reason for exclusion was selection bias – many studies only looked at targeted populations in their seroprevalence data, and thus could not be used as population estimators of IFR (11, 17-24). For some data, it was difficult to determine the numerator (i.e. number of deaths) associated with the seroprevalence estimate, or the denominator (i.e. population) was not well defined and thus we did not calculate an IFR (25, 26). One study explicitly warned against using its data to obtain an IFR (27). Another study calculated an IFR, but did not allow for an estimate of confidence bounds and thus could not be included in the quantitative synthesis (28).

After screening titles and abstracts, 227 studies were removed. Many of these looked at case-fatality estimates, or discussed IFR as a concept and/or a model input, rather than estimating the figure themselves. 40 papers were assessed for eligibility for inclusion into the study, resulting in a final 25 to be included in the qualitative synthesis.

Studies varied widely in design, with 3 entirely modelled estimates (29-31), 4 observational studies (10, 32-34), 5 pre-prints that were challenging to otherwise classify (2, 35-38), and a number of serological surveys of varying types reported by government agencies (39-51). For the purposes of this research, an estimate for New York City was calculated from official statistics and the serosurvey, however this was correlated with a published estimate (28) to ensure validity.

The main result from the random-effects meta-analysis is presented in Figure 1. Overall, the aggregated estimate across all 24 studies indicated an IFR of 0.68% (95% CI 0.53-0.82%), or 68 deaths per 10,000 infections. Heterogeneity was extremely high, with the overall I2 exceeding 99% (p<0.0001).

**Figure 1.**
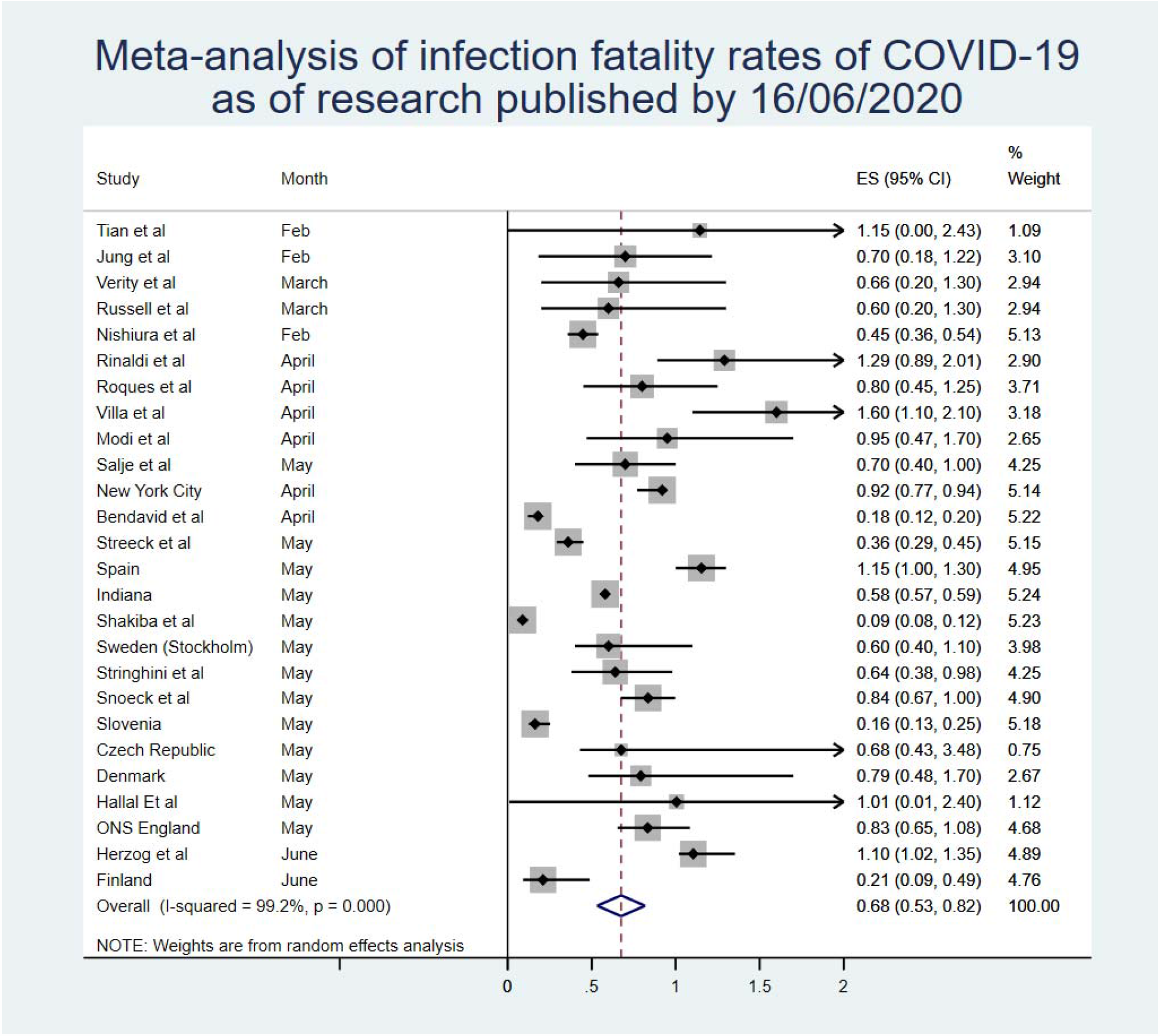

The sensitivity analysis by month from Figure 3 showed that earlier estimates of IFR were lower, with later estimates showing a higher figure, although this appears to have stabilised in May.

**Figure 2.**
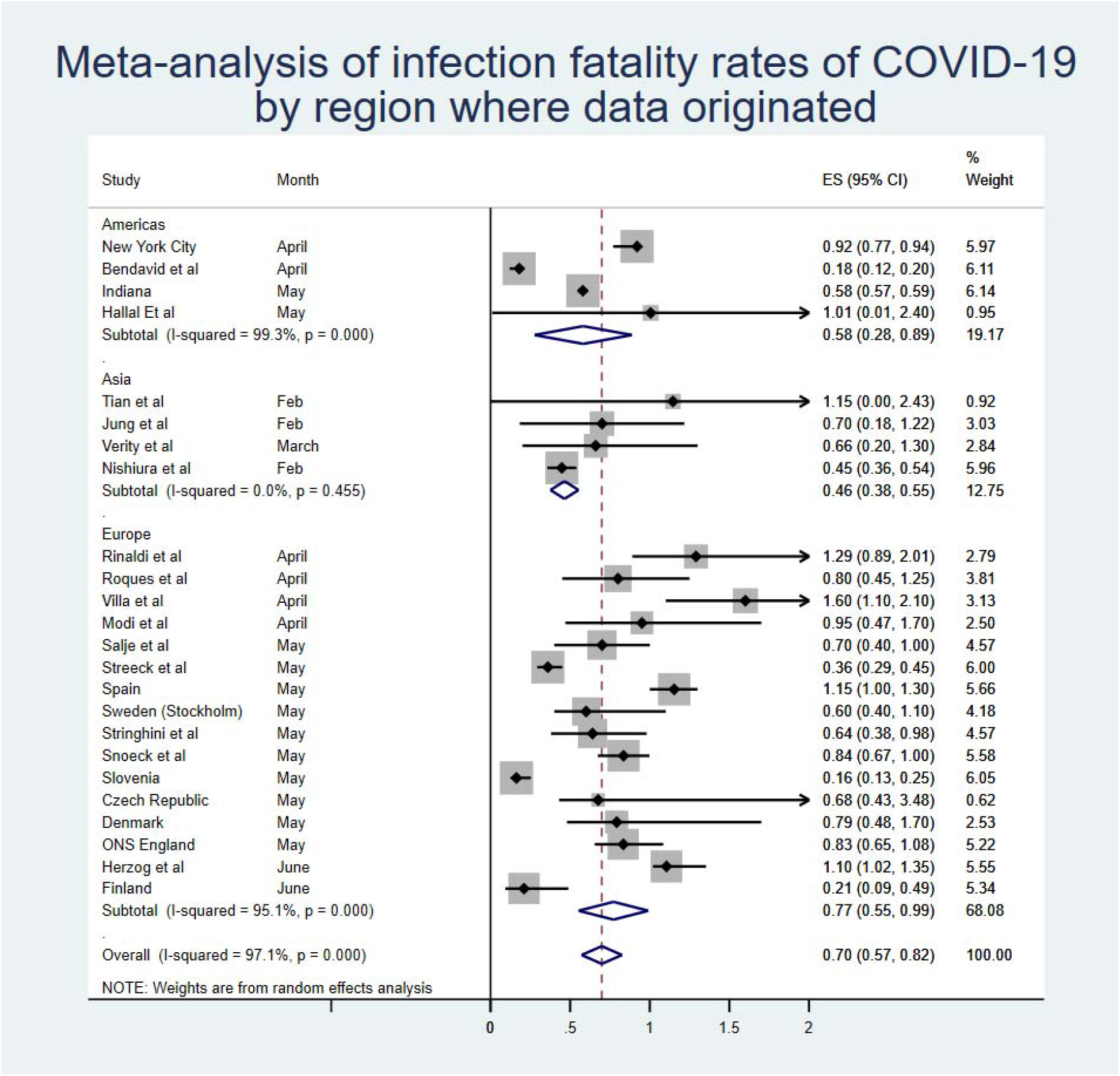

**Figure 3.**
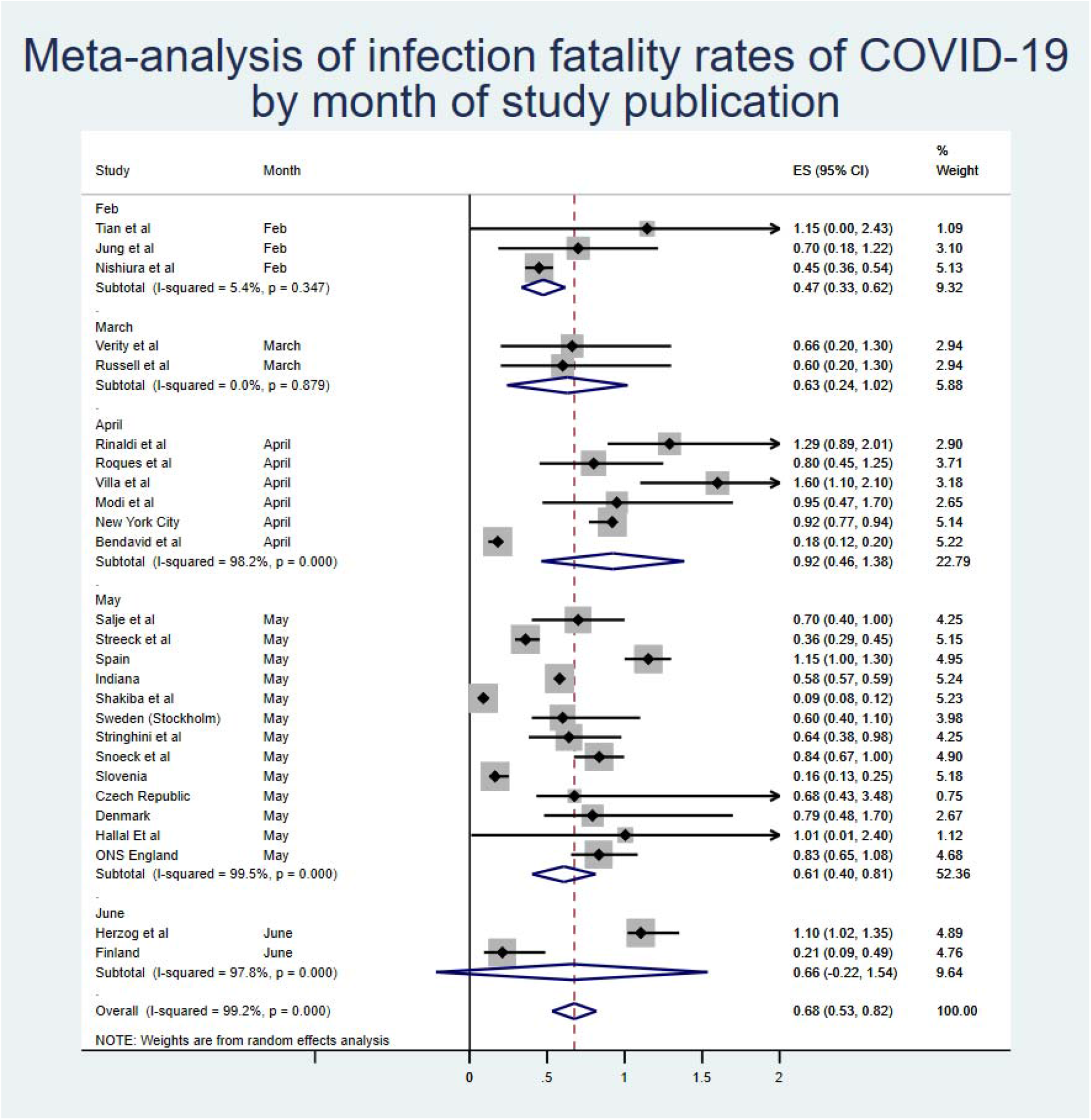

Analysing by region of origin did not appear to have a substantial effect on the findings, although there was a slightly lower estimate seen in Asia. As the Middle East was only represented by one study, this region was excluded from the meta-synthesis by region. Two studies were also excluded as they did not present an IFR for a specific region (i.e. Diamond Princess).

Of note, there was some difference in estimates of IFR between estimates based on serosurveys and those of modelled or PCR-based estimates. The overall estimates from serosurvey studies was 0.60% (0.42-0.77%), although again with very high heterogeneity, as can be seen in Figure 4.

**Figure 4.**
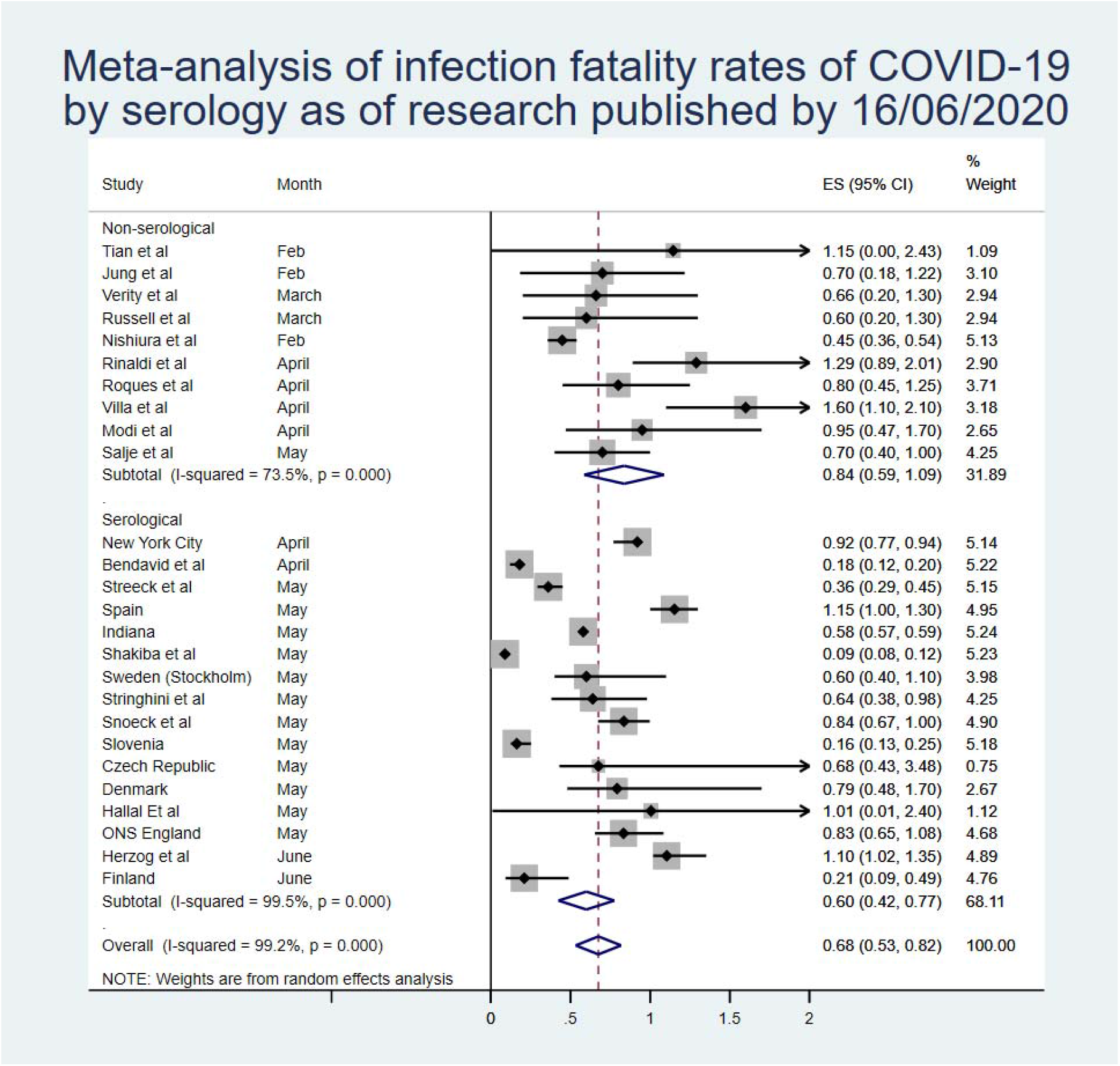

**Figure 5.**
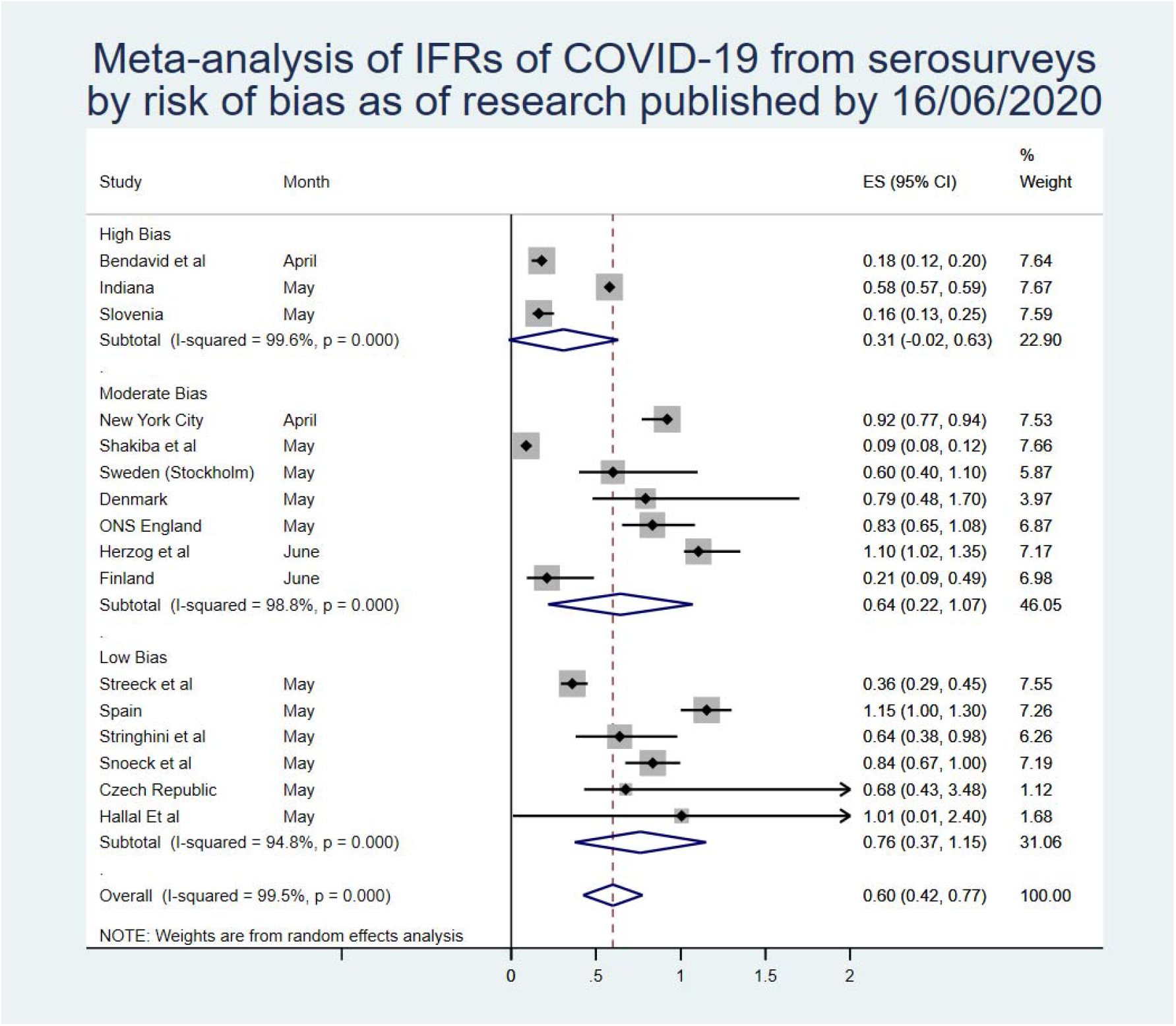

There were not sufficient data in the included research to perform a meta-analysis of IFR by age. However, qualitatively synthesizing the data that was presented indicates that the expected IFR below the age of 60 years is likely to be reduced by a large factor. This is supported by studies examining the CFR which were not included in the quantitative synthesis, as well as studies examining IFR in selected populations younger than 70 years of age, that demonstrate a strong age-related gradient to the death rate from COVID-19.

Plotting the studies using a funnel plot produced some visual indication of publication bias, with more high estimates than would be expected, however the Egger’s regression was not significant (p=0.74).

### Risk of Bias

As previously noted, all estimates obtained from modelling studies are considered to be at a high risk of bias due to the heterogeneity and difficulty in rating these studies for accuracy. After using the rating “risk of bias” tool for prevalence studies, 6 studies were considered to be at a low risk of bias, 4 studies at a moderate risk of bias, and the remaining 5 estimates at a high risk of bias. This is summarized in the table below (full scoring in supplementary materials):

In general, the primary reason for down-rating studies was non-response bias, the lack of representativeness of the population sample, and a lack of information across all fields. Some reports were published with minimal information, which substantially increased the uncertainty and thus the risk of bias in these estimates.

The sensitivity analysis by study quality results are below. Broadly, study quality was correlated with a higher inferred IFR, with lower-quality serosurveys reporting higher estimates of population prevalence than randomly-sampled population-wide prevalence estimates. Restricting the analysis to only those studies at a low risk of bias resulted in modestly reduced heterogeneity and an increased IFR of 0.76% (0.37-1.15%).

### Other Estimates of IFR

Several estimates of IFR were identified but not included in the meta-analysis as they did not meet the inclusion criteria. The aggregated best estimate from the Centre for Evidence-Based Medicine at Oxford University of 0.1-0.41% (52), and the preprint estimate reported by Grewelle and Leo of 1.04% (0.77-1.38%) (53) were both pertinent but could not be included due to collinearity.

Similarly, the estimate of symptomatic IFR produced by Basu of 1.3% (0.6-2.1%) (54) was excluded due to the exclusion of asymptomatic cases. Using reported estimates of asymptomatic cases, this estimate would likely match the meta-analytic IFR, however this correction could not be applied for the estimates in this study as it could easily introduce bias into the results.

## Discussion

As pandemic COVID-19 progresses, it is useful to use the IFR when reporting figures, particularly as some countries begin to engage in enhanced screening and surveillance, and observe an increase in positive cases who are asymptomatic and/or mild enough that they have so far avoided testing (55). It has been acknowledged that there is significant asymptomatic carriage – potentially up to 50% of all patients − and that asymptomatic transmission may also be possible with COVID-19 (29, 56) and use of IFR would aid the capture of these individuals in mortality figures. IFR modelling, calculation and figures, however, are inconsistent.

The main finding of this research is that there is very high heterogeneity among estimates of IFR for COVID-19 and therefore it is difficult to draw a single conclusion regarding the number. Aggregating the results together provides a point-estimate of 0.68% (0.53-0.82%), but there remains considerable uncertainty about whether this is a reasonable figure or simply a best guess. It appears likely, however, that the true population IFR in most places from COVID-19 will lie somewhere between the lower bound and upper bounds of this estimate.

One reason for the very high heterogeneity is likely that different countries and regions will experience different death rates due to the disease. One factor that may impact this is government response, with more prepared countries suffering lower death rates than those that have sufficient resources to combat a large outbreak (57). Moreover, it is very likely, given the evidence around age-related fatality, that a country with a significantly younger population would see fewer deaths on average than one with a far older population, given similar levels of healthcare provision between the two. For example, Israel, with a median age of 30 years, would expect a lower IFR than Italy, with a much higher median age (45.4 years).

Some included studies (2, 37) compared fatality during COVID-19 pandemic with previous years’ average fatality, determining that mortality has been higher during pandemic and whilst correlation doesn’t necessarily equate to causation, it is reasonable to link the events as causal given the high CFR observed across countries. It is highly likely from the data analysed that IFR increases with age-group, with those aged over 60 years old potentially experiencing the highest IFR, in one case close to 15% (37). Given the elderly are the most vulnerable in society to illness and likely to carry a higher disease burden owing to increased susceptibility and comorbidity (58, 59), the lower IFRs observed in the younger populations may skew the figure somewhat. There are some reasonable estimates of fatality in younger age-groups that were not included in the population estimates (11, 20, 23), which imply a substantially lower rate of death in the population under 70. While these studies were not considered applicable for quantitative synthesis, they imply that the IFR for <70 year olds is likely lower than 0.1%, and may be less than 10x the rate of death in over-70s. Another recently published estimate stratified infection-fatality by age and found a very low risk for under 50s that increased exponentially with age from 0.0016% <50 years to 0.14% for 50-64 year olds and up to 5.6% for those 65 years and older (60).

While not included in the quantitative synthesis, one paper did examine the extreme lower bound of IFR of COVID-19 in situations where the healthcare system has been overwhelmed. This is likely to be higher than the IFR in a less problematic situation but demonstrates that the absolute minimum in such a situation cannot be lower than 0.2%, and is likely much higher than this figure in most scenarios involving overburdened hospitals.

Of note, there appears to be a divergence between estimates based on serosurveys and those that are modelled or inferred from other forms of testing, with the IFR based purely on serosurveillance being 0.60% (0.43-0.77%). Some have argued that serological surveys are the only proper way to estimate IFR, which would lead to the acceptance of this slightly lower IFR as the most likely estimate (61). However, even these estimates are very heterogeneous in quality, with some extremely robust data such as that reported from the Spanish and Swedish health agencies (40, 62), and some that have clear and worrying flaws such as a study from Iran where death estimates are reportedly substantially lower than the true figure (45). However, when taking quality into account, and only analysing those serosurveys that had a low risk of bias, it is interesting to note that the inferred IFR rises substantially to 0.76% (0.37-1.15%). This may be due to the bias in lower-quality serosurveys being towards a higher prevalence (27), which in turn lowers the IFR substantially.

Another key issue is accounting for deaths. While official death counts were used for all serosurvey estimates, and included in all modelled estimates, these counts are increasingly being recognized as undercounts of the true death figure (37). Published research is already estimating that, even in many wealthy countries with excellent death reporting systems, more than 50% of COVID-19 deaths are likely being missed (63, 64). It is not unlikely that, after correcting for excess mortality not captured in official death reporting systems, the IFR of COVID-19 in most populations would be higher than 1%.

Conversely, there is evidence that the tests used in these serosurveys have drawbacks despite their high specificity and sensitivity. For example, in asymptomatic/mild cases, the tests may have reduced sensitivity, leading to a biased overestimation of the IFR (65). A recent systematic review and meta-analysis of serological tests for COVID-19 found that even the better serology tests would likely overestimate prevalence in an area with few cases and underestimate prevalence when many people had already been infected (9). In areas with a prevalence of 1-2%, for example, the systematic review implies that a study employing an enzyme-linked immunosorbent assay to examine antibodies would produce an estimated infection rate almost double the true prevalence. This would then cause the IFR be underestimated by the same fraction.

There are a number of limitations to this research. Importantly, the heterogeneity in the meta-analysis was very high. This may mean that the point-estimates are less reliable than would be expected. It is also notable that any meta-analysis is only as reliable as the data contained within – this research included a very broad range of studies that address slightly different questions with a very wide range of methodological rigor, and thus cannot represent certainty of any kind. While modelling studies were not formally graded, at least one has already been critiqued for simple mathematical errors, and given that many were pre-prints it is hard to ascertain if they have provided accurate representations of the data. Serology studies were at variable risk of bias, and analysing by only the highest quality serosurveys produced a higher estimate than relying on lower quality studies.

Moreover, the quality of included serosurvey estimates was often questionable. Many countries have a clear political motivation to present lower estimates, making it challenging to ascertain whether these may have biased the reporting of results, particularly for those places that have only presented results as press releases thus far. Some have also been criticized for sampling issues that would likely lead to a biased overestimate of population infection rates (10).

Accounting for right-censoring in these estimates was also a challenge. Using a 10-day cutoff for deaths is far too crude a method to create a reliable estimate. In some cases, this could be an overestimate, due to the seroconversion process taking almost as much time as the median time until death. Conversely, there is a long tail for COVID-19 deaths (15), and therefore it is almost certain that some proportion of the ‘true’ number of deaths will be missed by using a 10-day cutoff, biasing the estimated IFRs down. This may be why serosurvey estimates at first appear to result in somewhat lower IFRs than modelled and observational data suggests.

It is also important to recognize that this is a living estimate. With new data being published every single day during this pandemic, in a wide variety of languages and in innumerable formats, it is impossible to collate every single piece of information into one document no matter how rigorous. Moreover, this aggregated estimate is only as correct as the most recent search – the point estimate has not shifted substantially due to the inclusion of new research, but the confidence interval has changed. It is almost certain that, over the course of coming months and years, the IFR will be revised a number of times. In particular, it is vital that future research stratifies this estimate by age, as this appears to be the most significant factor in risk of death from COVID-19.

This research has a range of very important implications. Some countries have announced the aim of pursuing herd immunity with regards to COVID-19 in the absence of a vaccination. The aggregated IFR would suggest that, at a minimum, you would expect 0.45-0.53% of a population to die before the herd immunity threshold of the disease (based on R0 of 2.5-3 (34)) was reached (66). As an example, in the United States this would imply more than 1 million deaths at the lower end of the scale.

This also has implications for future planning. Governments looking to exit lockdowns should be prepared to see a relatively high IFR within the population who are infected, if COVID-19 re-emerges. This should inform the decision to relax restrictions, given that the IFR for people infected with COVID-19 appears to be not insignificant even in places with very robust healthcare systems.

## Conclusions

Based on a systematic review and meta-analysis of published evidence on COVID-19 until May, 2020, the IFR of the disease across populations is 0.68% (0.53-0.82%). However, due to very high heterogeneity in the meta-analysis, it is difficult to know if this represents the ‘true’ point estimate. In particular, higher-quality serosurveys with lower risk of bias appeared to generate higher IFRs. It is likely that, due to age and perhaps underlying comorbidities in the population, different places will experience different IFRs due to the disease. Given the issues with mortality recording, it is also likely that this represents an underestimate of the true IFR figure. More research looking at age-stratified IFR is urgently needed to inform policy-making on this front.

## Data Availability

Data and code available from authors on request

## Author declarations

The Authors declare no conflicts of interest. No funding was received for this study. A preprint version can be found here: https://www.medrxiv.org/content/10.1101/2020.05.03.20089854v1

This research was not funded.

